# Prognostic Association of Handgrip-Defined Probable or Possible Sarcopenia Status and Polygenic Risk with 10-Year Fracture Incidence among Black, Hispanic, and White Women: A Women’s Health Initiative Study

**DOI:** 10.64898/2026.06.30.26356931

**Authors:** Jongyun Jung, Qing Wu

**Author notes:** Correspondence: **Qing Wu, MD, ScD,** Department of Biomedical Informatics, College of Medicine, The Ohio State University Wexner Medical Center, 250 Lincoln Tower, 1800 Cannon Drive, Columbus, OH 43210.

## Abstract

**Purpose:** The Fracture Risk Assessment Tool (FRAX) excludes objective skeletal muscle health and genetic variables. We evaluated the prognostic associations of handgrip-defined probable/possible sarcopenia and genome-wide polygenic scores (GPS) with 10-year fracture risk, and their incremental predictive value beyond FRAX across racial/ethnic groups and GPS strata.

**Methods:** We analyzed 2,051 postmenopausal women from the Women’s Health Initiative. Race-specific analyses focused on Black, Hispanic, and White participants (n=2,009), excluding American Indian/Alaska Native and Asian/Pacific Islander individuals due to sparse fracture events. Sarcopenia status was operationalized by low handgrip strength alone via EWGSOP2 (<16.0 kg) and AWGS 2025 (<18.0–20.0 kg) criteria. Fine-Gray models estimated subdistribution hazard ratios (sHR), treating death as a competing risk. Predictive performance at 10 years was assessed using time-dependent AUC, Brier scores, and decision curve analysis (DCA).

**Results:** Handgrip-defined probable or possible sarcopenia prevalence was 4.4% (EWGSOP2) and 6.4% (AWGS 2025). Black women demonstrated lower risk for major osteoporotic fractures (MOF) (adjusted sHR=0.19, 95% CI: 0.08–0.48) and hip fractures (adjusted sHR=0.07, 95% CI: 0.01–0.52) compared to White women. Neither sarcopenia status nor high GPS showed statistically significant independent associations with fractures after FRAX adjustment. Adding sarcopenia status to baseline FRAX (AUC: 0.71 for MOF; 0.69 for hip) yielded near-identical AUCs, Brier scores, and within-sample net benefit.

**Conclusion:** Handgrip-defined probable/possible sarcopenia and current GPS do not provide independent or incremental predictive value beyond the clinical FRAX framework within this genomic sub-sample of older women.

**Mini Abstract:** The Fracture Risk Assessment Tool (FRAX) currently excludes skeletal muscle and genomic metrics. In 2,051 postmenopausal women, incorporating handgrip-defined sarcopenia and polygenic scores provided no incremental predictive benefit beyond standard FRAX. Within this study population, separate integration of these markers may not enhance routine fracture risk stratification.

## Introduction

Osteoporotic fractures represent a critical public health challenge, leading to increased morbidity, mortality, and substantial healthcare costs among postmenopausal women globally [1]. Currently, the Fracture Risk Assessment Tool (FRAX) serves as the international gold standard for predicting 10-year fracture probability [2,3]. However, the current FRAX tool relies primarily on clinical risk factors and bone mineral density (BMD), notably excluding objective measures of skeletal muscle health. This exclusion is significant because muscle health is biologically and mechanically intertwined with bone integrity, forming a “muscle-bone unit” where the decline of one often mirrors the other [4–6].

Sarcopenia is the age-related loss of muscle strength, quantity, and physical performance and linked to skeletal fragility and increased fracture risk [7,8]. Low grip strength and impaired physical function are robust, independent predictors of falls, which precede the vast majority of osteoporotic fractures [4–6]. Despite the clear biological link, it remains unclear whether sarcopenia provides independent predictive value once traditional clinical risk factors are already accounted for within the FRAX framework. This study focuses on EWGSOP2 “probable sarcopenia” [7] and AWGS 2025 “possible sarcopenia” [9]. EWGSOP2 elevates low muscle strength as the primary indicator, noting it is sufficient to trigger clinical assessment and intervention. Similarly, AWGS 2025 emphasizes possible sarcopenia as a critical threshold for early detection in community settings. Although AWGS 2025 permits low strength or poor physical performance, we operationalize both frameworks exclusively through low handgrip strength to ensure objective, consistent muscle health assessment [9].

Two major gaps persist in the current literature that limit our understanding of this relationship. First, although significant disparities in fracture incidence exist across racial and ethnic groups [10–12], no studies to date have systematically examined whether the addition of probable or possible sarcopenia to the FRAX model improves predictive accuracy across the Black, Hispanic, and White racial and ethnic populations. Universal probable or possible sarcopenia thresholds may not provide equitable risk assessment, given that muscle mass and strength vary by ethnicity [13,14]. Second, while genome-wide polygenic scores (GPS) identify genetic susceptibility to fractures [15–17], no study has examined if probable or possible sarcopenia provides additive predictive value in individuals across different genomic risk strata. It is currently unknown whether probable or possible sarcopenia provides additive predictive information in individuals already categorized into different genomic risk strata.

The objective of this study was to evaluate the prognostic associations of probable or possible sarcopenia and GPS with fracture risk and to determine the incremental predictive value of incorporating sarcopenia status into the FRAX framework. Specifically, we aimed to evaluate whether the prognostic association of sarcopenia persists across different racial/ethnic groups and across high, medium, and low GPS strata.

## Materials and Methods

### Study Design

This study is a retrospective observational cohort analysis utilizing data from the Women’s Health Initiative (WHI), a national, longitudinal research program addressing prevalent health issues such as cardiovascular disease, cancer, and osteoporosis among postmenopausal women aged 50–79 years [18]. This study adheres to the Strengthening the Reporting of Observational Studies in Epidemiology (STROBE) guidelines [19].

### Participants

Participants included postmenopausal women enrolled in six WHI sub-studies, namely: Genomics and Randomized Trials Network (GARNET, n=4,894), SNP Health Association Resource (SHARE, n=12,007), Population Architecture using Genomics and Epidemiology (PAGE, n=12,646), Women’s Health Initiative Memory Study (WHIMS, n=5,740), WHI Sequencing Project (WHISP, n=1,904), and Genome-Wide Association Study (GWAS) Hip Fracture (n=3,688). After merging these sub-studies, an initial total of 40,879 participants was obtained. Participants lacking race/ethnicity data (n=2,237), individuals with overlapping participation in multiple studies (n=10,634), those on osteoporosis medications (e.g., bisphosphonates, calcitonin, selective estrogen receptor modulators, parathyroid hormone) (n=405), and those self-identifying as ‘Other’ (n=91) were systematically excluded, yielding a final analytical sample of 27,512 unique participants. From this cohort, we specifically selected 2,051 participants who possessed complete baseline data for muscle strength and genetic information (**Figure 1**, **Table S1**). This final sample was composed of participants originally derived from WHIMS (n=733), GARNET (n=501), PAGE (n=345), the GWAS Hip Fracture study (n=230), SHARE (n=177), and WHISP (n=65).

**Figure 1.**
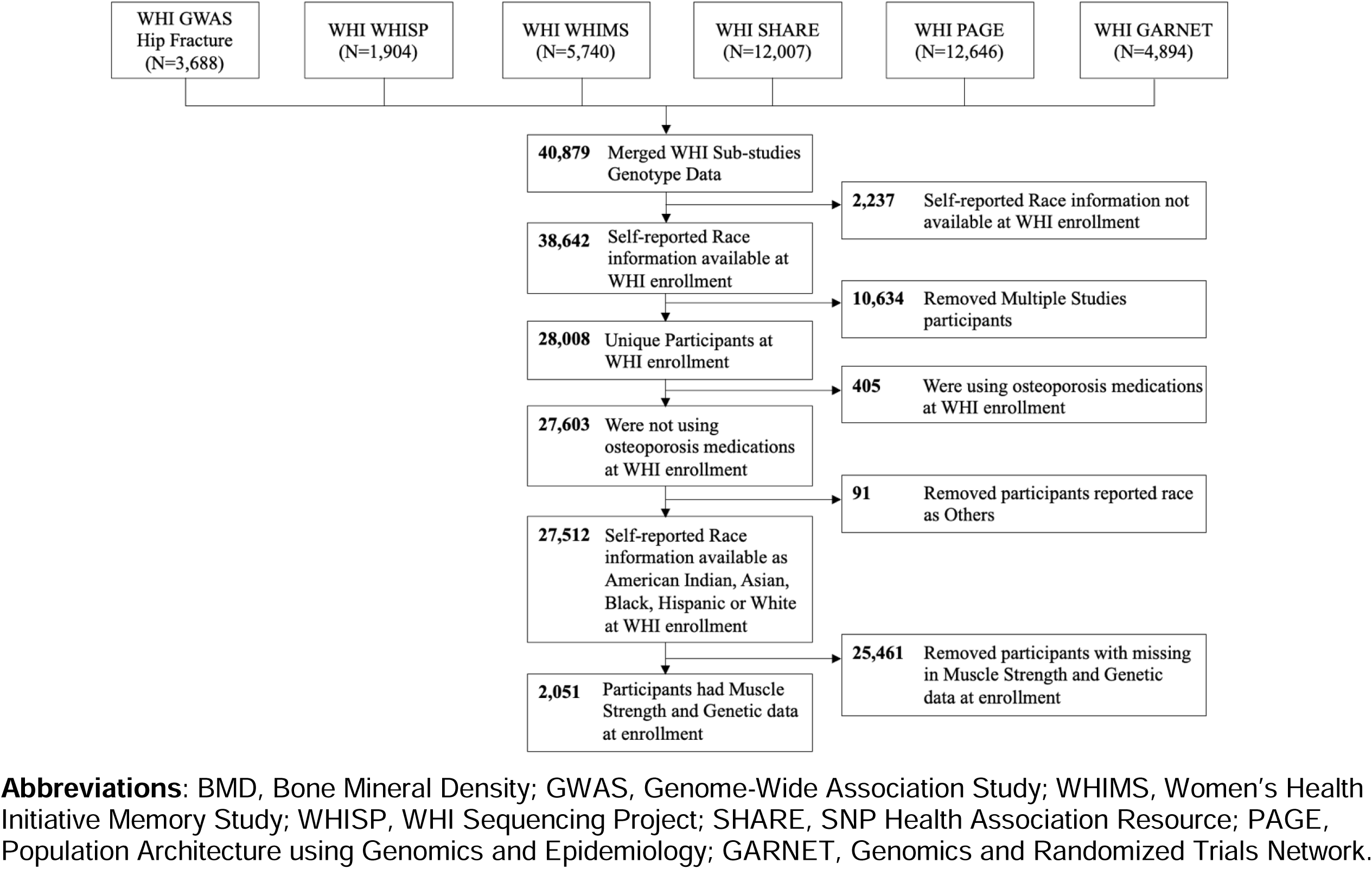
Study Flow Diagram for the Derivation of the Analytical Cohort from Women’s Health Initiative (WHI) Participants.

To evaluate potential selection bias, we compared baseline characteristics between the final analytic cohort (n=2,051) and those excluded (n=25,461) (**Table S2**). The analytic cohort was older (70.1 vs. 63.2 years, SMD: 1.17) with a higher prevalence of White participants (73.1% vs. 40.2%, SMD: 0.72). The 10-year cumulative incidence of MOF (6.5% vs. 5.1%, SMD: 0.06) and hip fractures (3.6% vs. 3.3%, SMD: 0.01) did not meaningfully differ. Baseline FRAX scores (without BMD) for MOF (9.6% vs. 8.2%, SMD: 0.08) and hip fractures (2.7% vs. 2.1%, SMD: 0.05) remained comparable. This suggests that despite representing an older phenotype with minor risk variations, the analytic cohort’s clinical risk profiles remain consistent with the broader WHI population.

### Ethical Considerations

All WHI study protocols received approval from Institutional Review Boards at all participating institutions [20]. Genomic data access was granted by the database of Genotype and Phenotype (dbGaP) [21] with institutional approval from Ohio State University (IRB approval: STUDY20250826). Written informed consent, including explicit permission for genetic analyses and DNA usage, was obtained from all WHI participants.

### Muscle Measurement and Assessment

Muscle strength was assessed via handgrip strength using a calibrated Jamar hydraulic dynamometer (Sammons Preston Rolyan, Bolingbrook, IL). Participants performed two maximal trials per hand while seated with the elbow flexed at 90°. The maximum value from either hand was recorded and utilized as the primary strength metric for the diagnosis of sarcopenia.

### Sarcopenia Definitions

For the primary analysis, probable or possible sarcopenia risk was characterized using two clinical consensus frameworks: the European Working Group on Sarcopenia in Older People 2 (EWGSOP2) [7] and the Asian Working Group for Sarcopenia 2025 Consensus Update (AWGS 2025) [9].

Following the clinical pathways outlined by both groups, we focused on the earliest clinically actionable stages: “probable sarcopenia” (EWGSOP2) and “possible sarcopenia” (AWGS 2025). The EWGSOP2 framework identifies “probable sarcopenia” based on low muscle strength alone, noting that this is a sufficient threshold to trigger clinical assessment and intervention [7]. Similarly, the AWGS 2025 consensus emphasizes “possible sarcopenia”, defined by low muscle strength or poor physical performance, as the critical stage for early detection in community settings [9].

Under the EWGSOP2 framework, probable sarcopenia was defined as a maximum handgrip strength < 16.0 kg. To maintain consistency with the grip strength-based EWGSOP2 operationalization, possible sarcopenia under the AWGS 2025 framework was defined in this study with low muscle strength only using age-specific thresholds: < 20.0 kg for participants aged 50–64 years and < 18.0 kg for those 65 years and older. These binary classifications (Normal vs. Probable or Possible Sarcopenia) served as the primary exposure variables for all fracture risk models. Because grip strength alone was utilized in this study, we therefore use the term ‘handgrip-defined probable or possible sarcopenia throughout in this study.

### Genetic Analysis and Genome-Wide Polygenic Scores

Genotyping was conducted using Illumina (Illumina Inc., San Diego, CA, USA) or Affymetrix 6.0 Array platforms (Affymetrix Inc., Santa Clara, CA, USA). Rigorous quality control included filtering for minor allele frequency (MAF ≥ 0.01), individual missingness (<5%), and deviation from Hardy-Weinberg equilibrium (P < 0.0001) using PLINK 1.9 [22] . Genotype imputation was performed via the Michigan Imputation Server [23] using the Haplotype Reference Consortium panel [24] and the 1000 Genomes Phase 3 (Version 5) [25] to ensure genomic representation across diverse populations. The GPS for fracture risk was computed employing the LDpred Bayesian algorithm [26], utilizing summary statistics from a large-scale UK Biobank genome-wide association study involving 103,155 genetic variants [17]. To account for population stratification and technical variability, all models involving the GPS were adjusted for the genotyping array, sub-study indicators, and the first 10 genetic principal components. GPS was categorized into Low (bottom 5%), Medium (middle 90%), and High (top 5%) risk groups within ancestry strata to ensure standardized representation across the analytic cohort.

### Fracture Outcomes and Follow-Up

The primary outcome was the occurrence of MOF, defined as fractures of the hip, clinical spine, forearm, or shoulder. Hip fracture was the secondary outcome. Fractures were identified through standardized self-reported questionnaires administered annually or semiannually and confirmed by centralized adjudication using radiologic or operative reports reviewed by blinded adjudicators [27,28]. The index date (baseline) for this study was defined as the date of enrollment, at which time participants provided complete baseline measurements for muscle strength (grip strength) and genetic data. Participants were followed for up to 10 years or until the first fracture event or death, with deaths treated as competing risks. Participants lost to follow-up were censored at their last known contact. For hip-fracture models, follow-up was censored at the first non-hip MOF event; these results should be interpreted as first-fracture analyses rather than definitive hip-fracture incidence estimates. Time zero was WHI baseline enrollment; grip strength measurements, FRAX covariates, and genotype-derived GPS eligibility were all ascertained at this date. Follow-up was administratively truncated at 10 years to align with the FRAX prediction horizon.

### Covariates and Clinical Risk Factors

Covariates included demographic (age, self-reported race/ethnicity), anthropometric (height, weight), lifestyle (smoking status, alcohol intake), and clinical factors (parental hip fracture history, previous fractures, rheumatoid arthritis, ≥ 1 Previous falls in the past 12 months). These were collected through standardized questionnaires and clinical assessments. Smoking status was categorized as “Never”, “Past”, or “Current”. Fall history was dichotomized based on self-reported falls within the preceding 12 months.

### FRAX Risk Assessment

The 10-year probabilities for MOF and hip fracture were assessed using the US FRAX tool (version 3.0). To ensure full transparency and reproducibility, this study utilized pre-computed baseline FRAX scores provided by the WHI clinical coordinating center (variables WHOFRAC for MOF and WHOHIP for hip fracture), which were retrieved from the dbGaP. Crucially, these scores were calculated without the incorporation of BMD values to evaluate the incremental predictive value of sarcopenia and genetic risk over the standard clinical FRAX algorithm.

### Statistical Analysis

The final analytic sample consisted of 2,051 participants with complete data for muscle strength and genotypes at the enrollment index date. To address minor data gaps while preserving the cohort, multiple imputation using predictive means matching was employed for continuous variables with negligible missingness, including age (n=2), weight (n=5), and height (n=3). The baseline burden of probable or possible sarcopenia and fracture risk was characterized by calculating the prevalence of EWGSOP2 and AWGS 2025 criteria across study strata, with the cumulative incidence of fractures and the competing risk of mortality estimated using the Aalen-Johansen cumulative incidence function (CIF).

Baseline characteristics were compared between participants with and without probable or possible sarcopenia. P-values were obtained using independent t-tests for continuous variables and chi-square tests (or Fisher’s exact tests where appropriate) for categorical variables. To evaluate the magnitude of imbalance between groups independent of sample size, standardized mean differences (SMD) were calculated. An SMD > 0.1 was considered a small imbalance, while an SMD > 0.2 represented a meaningful imbalance.

For each fracture outcome, we developed a series of Fine-Gray subdistribution hazard models to estimate subdistribution hazard ratios (sHR) and 95% confidence intervals (CI), treating death as a competing risk [29]. The sHR represents the relative change in the subdistribution hazard of the fracture event associated with a unit change in the predictor, accounting for the fact that participants who die are no longer at risk for the event but remain in the denominator [29]. To evaluate the prognostic association of sarcopenia status with 10-year fracture risk, we constructed three sequential models:

- Model 1 (Base Model): Evaluated the relationship between the outcome and the continuous FRAX score (without BMD) alone.
- Model 2 (Sarcopenia-Augmented Model): Included the FRAX score plus binary sarcopenia status (Normal vs. Probable or Possible) to determine if muscle strength adds incremental prognostic value to established clinical risk scores.
- Model 3 (Covariate-Adjusted Sensitivity Model): Included probable or possible sarcopenia status, age, height, and weight as independent predictors. This model was designed to assess the association of muscle strength with fractures independent of the specific internal weighting and clinical variables utilized within the FRAX algorithm.

To evaluate whether the association between sarcopenia and fracture risk varied by demographic or genetic factors, formal interaction terms (Race × Sarcopenia and GPS Sarcopenia) were tested using the Fine-Gray subdistribution hazard framework. Interaction p-values were derived from likelihood ratio tests comparing models with and without the interaction terms. For the race-stratified analysis (n = 2,009), small-sample groups (American Indian/Alaska Native and Asian/Pacific Islander) were excluded to maintain model stability.

The incremental predictive value of adding probable or possible sarcopenia to the baseline FRAX model was assessed at a 10-year horizon. This study is a model-update and internal validation analysis; we specifically evaluated the FRAX framework (apparent validation) and its extension via muscle health indicators (model development/update).

Discrimination was evaluated via the time-dependent Area Under the Curve (AUC) using Inverse Probability of Censoring Weighting (IPCW), while overall accuracy was measured by the Brier Score. Specifically, time-dependent AUC was estimated using the IPCW concordance statistic for competing-risk outcomes (R package timeROC). Brier scores represent the mean squared difference between predicted 10-year CIF and the observed cause-specific outcome indicator, with death treated as a competing event (R package riskRegression). Calibration was assessed by comparing mean predicted 10-year CIF against observed CIF within deciles of predicted risk (calibration-in-the-large and calibration slope). DCA used the net-benefit formulation adapted for competing risk, with death as the competing event (R package dcurves). Key R packages: survival, cmprsk, riskRegression, timeROC, dcurves, mice.

Model calibration was evaluated using Calibration-in-the-large (CITL) and calibration slope. To account for potential over-fitting and to provide realistic performance estimates, we employed a bootstrap optimism-correction procedure with 1,000 resamples [30]. Optimism was calculated as the average difference in performance between the bootstrap-derived model tested on the bootstrap sample versus the original dataset. All reported metrics (AUC, Brier, CITL, and Slope) are optimism-corrected.

Decision Curve Analysis (DCA) was implemented to determine the net clinical benefit. To quantify the incremental value of probable or possible sarcopenia, we calculated the change (Δ) in AUC, Brier Score, and Net Benefit compared to the baseline FRAX model. The decision context for net benefit was defined by the clinically relevant 10-year fracture risk thresholds of 20% for MOF and 3% for Hip fracture, representing the standard cut-offs for pharmacological intervention. To ensure reproducibility, we reported the model coefficients and the 10-year baseline cumulative incidence (F_0_(t)) for the augmented models to allow for external geographic or temporal validation.

### Sampling and Generalizability

To maximize statistical power for stratified analyses, we utilized the full analytic sample (n=2,051). Because our cohort comprises multiple sub-studies with varying designs, including the case-control GARNET study (n=501) and the age-restricted WHIMS cohort (n=733), we did not apply sampling weights. Consequently, all cumulative incidence, calibration, and net benefit metrics reflect within-sample performance. These outputs evaluate the internal validity of the sarcopenia and genomic markers within this specific cohort; they are non-transportable to the general postmenopausal population without external validation in prospective, population-based samples.

### Sensitivity Analyses

To maintain maximal analytic power for the covariate-adjusted models (n=2,051), multiple imputation with predictive means matching was used to address negligible missingness in baseline physical characteristics. Specifically, two values for age, three for height, and five for weight were imputed. As described in the statistical modeling section, these imputed values were utilized exclusively in Model 3 to ensure the robustness of the observed racial and genetic associations. This sensitivity model assessed the prognostic association between probable or possible sarcopenia and fracture risk (sHR), controlling for age, height, and weight independently of the internal variable weighting within the standard FRAX algorithm.

### Software and Statistical Significance

Statistical analyses were performed using R version 4.4.0 (The R Foundation). A two-tailed P-value of <0.05 was considered statistically significant throughout analyses.

## Results

### Baseline Characteristics and Sarcopenia Prevalence

The final analytic cohort (n=2,051) was characterized by an older mean age (70.1 years) and a predominantly White population (73.1%). Baseline characteristics stratified by EWGSOP2 probable sarcopenia are detailed in **Table 1**. The overall prevalence of probable or possible sarcopenia was 4.4% (n=91) according to EWGSOP2 (**Table 1**) and 6.4% (n=132) using AWGS 2025 criteria (**Table S3**).

**Table 1.**
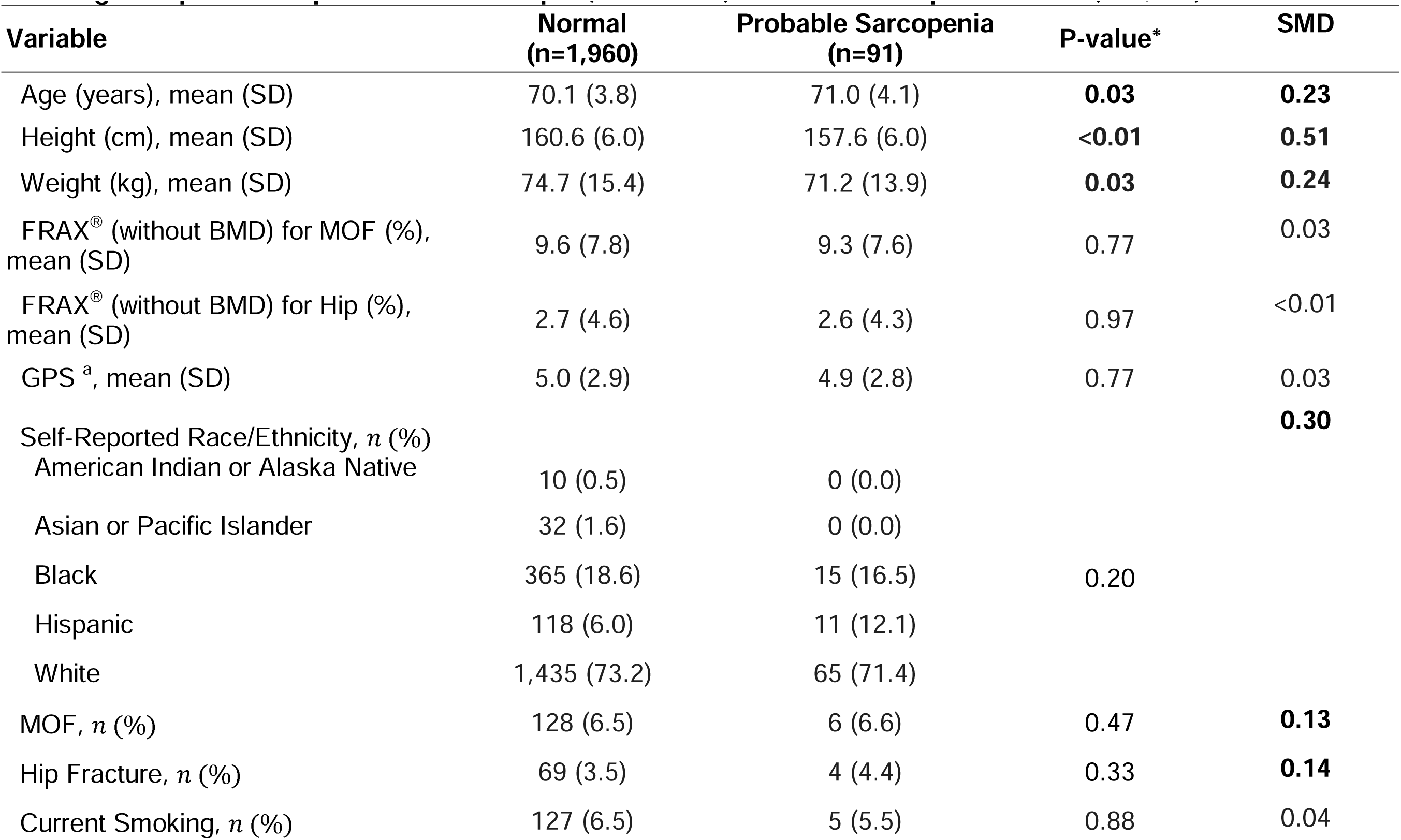
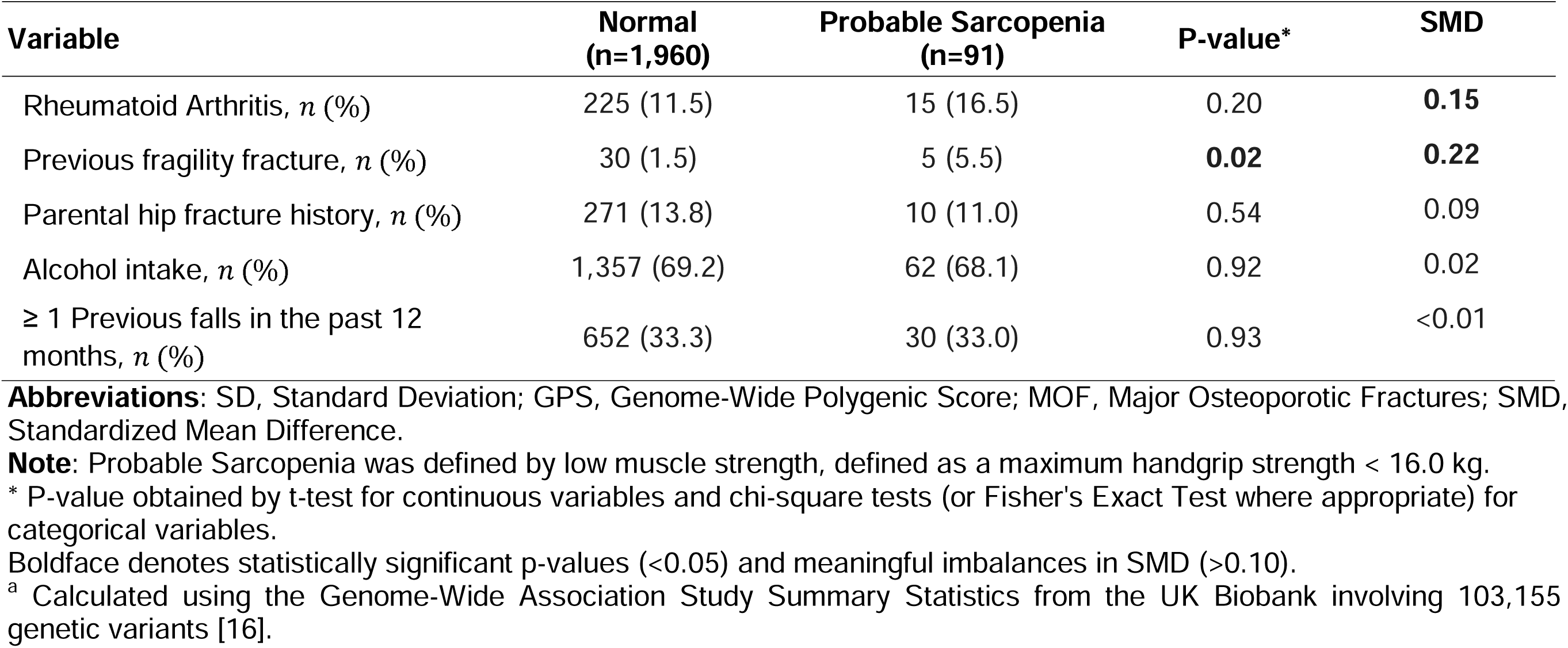
Baseline Characteristics of Participants in the Women’s Health Initiative Study Stratified by the European Working Group on Sarcopenia in Older People (EWGSOP2) Probable Sarcopenia Status (n=2,051).

| Variable | Normal<br>(n=1,960) | Probable Sarcopenia<br>(n=91) | P-value* | SMD |
| --- | --- | --- | --- | --- |
| Age (years), mean (SD) | 70.1 (3.8) | 71.0 (4.1) | **0.03** | **0.23** |
| Height (cm), mean (SD) | 160.6 (6.0) | 157.6 (6.0) | **<0.01** | **0.51** |
| Weight (kg), mean (SD) | 74.7 (15.4) | 71.2 (13.9) | **0.03** | **0.24** |
| FRAX<sup>®</sup> (without BMD) for MOF (%), mean (SD) | 9.6 (7.8) | 9.3 (7.6) | 0.77 | 0.03 |
| FRAX<sup>®</sup> (without BMD) for Hip (%), mean (SD) | 2.7 (4.6) | 2.6 (4.3) | 0.97 | <0.01 |
| GPS <sup>a</sup>, mean (SD) | 5.0 (2.9) | 4.9 (2.8) | 0.77 | 0.03 |
| Self-Reported Race/Ethnicity, *n* (%) |  |  |  | **0.30** |
| American Indian or Alaska Native | 10 (0.5) | 0 (0.0) |  |  |
| Asian or Pacific Islander | 32 (1.6) | 0 (0.0) |  |  |
| Black | 365 (18.6) | 15 (16.5) | 0.20 |  |
| Hispanic | 118 (6.0) | 11 (12.1) |  |  |
| White | 1,435 (73.2) | 65 (71.4) |  |  |
| MOF, *n* (%) | 128 (6.5) | 6 (6.6) | 0.47 | **0.13** |
| Hip Fracture, *n* (%) | 69 (3.5) | 4 (4.4) | 0.33 | **0.14** |
| Current Smoking, *n* (%) | 127 (6.5) | 5 (5.5) | 0.88 | 0.04 |
| Rheumatoid Arthritis, *n* (%) | 225 (11.5) | 15 (16.5) | 0.20 | **0.15** |
| Previous fragility fracture, *n* (%) | 30 (1.5) | 5 (5.5) | **0.02** | **0.22** |
| Parental hip fracture history, *n* (%) | 271 (13.8) | 10 (11.0) | 0.54 | 0.09 |
| Alcohol intake, *n* (%) | 1,357 (69.2) | 62 (68.1) | 0.92 | 0.02 |
| ≥ 1 Previous falls in the past 12 months, *n* (%) | 652 (33.3) | 30 (33.0) | 0.93 | <0.01 |

**Abbreviations:** SD, Standard Deviation; GPS, Genome-Wide Polygenic Score; MOF, Major Osteoporotic Fractures; SMD, Standardized Mean Difference.
**Note:** Probable Sarcopenia was defined by low muscle strength, defined as a maximum handgrip strength < 16.0 kg.
\* P-value obtained by t-test for continuous variables and chi-square tests (or Fisher's Exact Test where appropriate) for categorical variables.
Boldface denotes statistically significant p-values (<0.05) and meaningful imbalances in SMD (>0.10).
<sup>a</sup> Calculated using the Genome-Wide Association Study Summary Statistics from the UK Biobank involving 103,155 genetic variants [16].

Across both frameworks, participants with probable or possible sarcopenia exhibited substantial imbalances in physical characteristics compared to those with normal muscle status. Specifically, probable or possible sarcopenic individuals had meaningfully lower mean height (SMD: 0.51 for EWGSOP2; 0.43 for AWGS 2025) and weight (SMD: 0.24 for EWGSOP2; 0.31 for AWGS 2025). Meaningful imbalances were also observed for age (SMD: 0.23 for EWGSOP2) and previous fragility fractures (SMD: 0.22 for EWGSOP2; 0.21 for AWGS 2025).

Of the 2,051 participants, 42 individuals (American Indian/Alaska Native, n=10; Asian/Pacific Islander, n=32) were excluded from racial stratification and regression models due to a lack of fracture events, resulting in an analytical sample of 2,009 Black, Hispanic, and White women. When stratified by self-reported race/ethnicity (**Table S4**), prevalence varied across the major groups; Hispanic participants exhibited the highest rates (8.5% by EWGSOP2; 10.1% by AWGS 2025), while Black participants exhibited the lowest (3.9% by EWGSOP2; 4.2% by AWGS 2025). When stratified by genomic risk (**Table S5**), the prevalence of possible sarcopenia remained relatively stable across GPS categories. Specifically, the prevalence of AWGS 2025 possible sarcopenia was 6.8% in the High GPS group and 4.9% in the Low GPS group, with the Medium GPS group exhibiting a prevalence of 6.5%. For the EWGSOP2 criteria, prevalence rates were 3.9%, 4.6%, and 2.9% for the High, Medium, and Low GPS groups, respectively.

### Follow-up and Fracture Event Accrual

Participants were followed for a maximum duration of 10 years to align with the FRAX prediction horizon, resulting in a median follow-up of 8.7 years (IQR: 0.55–10.0). During this period, 134 (6.5%) participants experienced the MOF, and 73 (3.6%) participants experienced a hip fracture. A substantial proportion of the cohort (n = 335, 16.3%) experienced the competing risk of death prior to a fracture event within the 10-year window.

### Subgroup and Interaction Analyses

To determine whether the prognostic value of probable or possible sarcopenia status varied by Race/Ethnicity and GPS, we tested formal interaction terms within our age, height, and weight adjusted models. We observed no statistically significant interactions between Race/Ethnicity and Sarcopenia status for either MOF (P_interaction_ = 0.42 for EWGSOP2; P_interaction_ = 0.38 for AWGS 2025) or hip fracture (P_interaction_ = 0.88 for EWGSOP2; P_interaction_ = 0.91 for AWGS 2025). Similarly, the interactions between GPS strata and probable or possible sarcopenia status were not statistically significant for MOF (P_interaction_ = 0.55) or hip fracture (P_interaction_ = 0.72). No statistically significant interaction was detected; however, sparse events in several race/ethnicity and GPS strata, including strata with zero events in **Table S4** and **S5**, limit inference about effect heterogeneity. These findings should be considered hypothesis-generating only and do not exclude the possibility of clinically meaningful effect modifications.

### 10-Year Cumulative Incidence of Fractures and Probable or Possible Sarcopenia Status

The 10-year cumulative incidence of fracture events stratified by the probable or possible sarcopenia status is presented in **Table 2**. In the overall cohort, the probability of MOF among those with AWGS 2025 possible sarcopenia was 10.8% (95% CI: 5.7%–17.8%) compared to 8.8% (95% CI: 7.4%–10.4%) in the normal group. Conversely, under EWGSOP2 criteria, the 10-year incidence of MOF was similar between the probable sarcopenia (8.7%, 95% CI: 3.5%-16.9%) and normal (9.0%, 95% CI: 7.6%-10.5%) groups. For hip fractures, incidence was slightly higher in the EWGSOP2 probable sarcopenia group (6.0%, 95% CI: 1.9%-13.6%) compared to the normal group (5.1%, 95% CI: 4.0%-6.3%).

**Table 2.** 10-Year Cumulative Incidence of Major Osteoporotic and Hip Fractures Stratified by the Handgrip-Defined Probable or Possible Sarcopenia Status According to the European Working Group on Sarcopenia in Older People (EWGSOP2) and the Asian Working Group for Sarcopenia 2025 Consensus Update (AWGS 2025) Criteria (n=2,051).

| Fracture | Sarcopenia Status | EWGSOP2 |  |  | AWGS 2025 |  |  |
| --- | --- | --- | --- | --- | --- | --- | --- |
|  |  | n | Events | 10-Year Cumulative Incidence (95% CI) | n | Events | 10-Year Cumulative Incidence (95% CI) |
| MOF | Normal | 1,960 | 128 | 9.0% (7.6% - 10.5%) | 1,919 | 123 | 8.8% (7.4% - 10.4%) |
|  | Probable/Possible | 91 | 6 | 8.7% (3.5% - 16.9%) | 132 | 11 | 10.8% (5.7% - 17.8%) |
| Hip | Normal | 1,960 | 69 | 5.1% (4.0% - 6.3%) | 1,919 | 68 | 5.1% (4.0% - 6.4%) |
|  | Probable/Possible | 91 | 4 | 6.0% (1.9% - 13.6%) | 132 | 5 | 5.4% (2.0% - 11.3%) |

**Abbreviations:** MOF, Major Osteoporotic Fractures; Hip, Hip Fracture.
**Note:** The 10-Year Cumulative Incidence values represent the estimated 10-year probability of the fractures. These estimates were calculated using the Aalen-Johansen estimator to account for the competing risk of mortality and right-censoring. This approach ensures that the reported risk is not overestimated by ignoring those who died before experiencing a fracture. EWGSOP2 Probable Sarcopenia was defined by low muscle strength, defined as a maximum handgrip strength < 16.0 kg. AWGS 2025 Possible Sarcopenia was defined according to the AWGS 2025 consensus as the presence of low muscle strength (grip strength <20.0 kg for ages 50–64 years; <18.0 kg for ages ≥ 65 years).

### Probable or Possible Sarcopenia Status and Fracture Risk

In the total analytic cohort (n=2,051), Fine-Gray subdistribution hazard models were used to evaluate the association between probable or possible sarcopenia status and fracture hazards (**Table 3**). For MOF, the baseline FRAX Only model showed a significant subdistribution hazard ratio of 1.05 (95% CI: 1.03–1.06). When sarcopenia was added, AWGS 2025 possible sarcopenia exhibited higher point estimates for MOF risk in both the FRAX-adjusted (sHR = 1.22, 95% CI: 0.67–2.23) and covariate-adjusted (sHR = 1.24, 95% CI: 0.67–2.28) models, though these did not reach statistical significance. EWGSOP2 probable sarcopenia showed no increased hazard for MOF (Adjusted sHR = 0.99, 95% CI: 0.45-2.21). Regarding hip fractures, the FRAX Only model demonstrated a significant subdistribution hazard ratio of 1.06 (95% CI: 1.03–1.08). In the adjusted model (Model 3), individuals with EWGSOP2 probable sarcopenia showed a 21% increased hazard for hip fracture (sHR = 1.21, 95% CI: 0.52–3.82) compared to the normal group, while the AWGS 2025 possible group showed an 11% increased hazard (sHR = 1.11, 95% CI: 0.45–2.74).

**Table 3.**
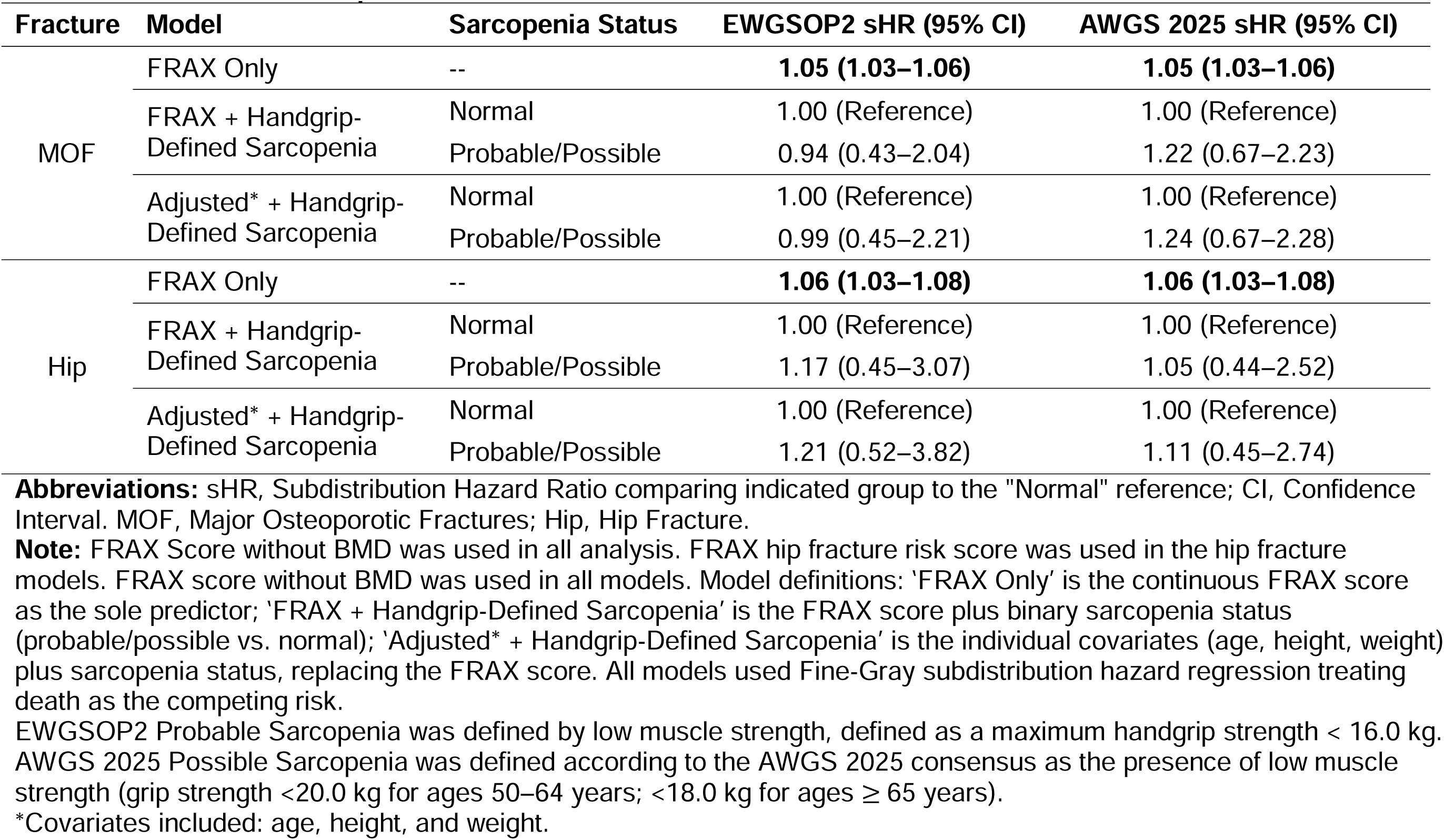
The Subdistribution Hazard Ratios (95% CI) for 10-Year Major Osteoporotic and Hip Fracture Risk by the Probable or Possible Sarcopenia Status (n=2,051).

### Race/Ethnicity, Genome-Wide Polygenic Score and Fracture Risk

Significant differences in fracture risk were observed by race/ethnicity (**Table S6**). Using White participants as the reference, Black women demonstrated lower risk for both MOF (Adjusted sHR = 0.19, 95% CI: 0.08–0.48) and hip fracture (Adjusted sHR = 0.07, 95% CI: 0.01–0.52). Hispanic women demonstrated a lower point estimate for hip fracture risk (sHR = 0.31; 95% CI: 0.04–2.19); however, this association was statistically non-significant and highly imprecise, as evidenced by a wide confidence interval that spans the null value of 1.00.

Beyond demographic strata, we examined the prognostic association of genomic risk using GPS categories (**Table S7**). We observed a positive trend where higher genomic risk corresponded with elevated point estimates for fracture risk. Specifically, participants in the High GPS group exhibited a more than two-fold increase in the subdistribution hazard for MOF (Adjusted sHR = 2.34; 95% CI: 0.81–6.81) compared to the Low GPS group. However, these associations remained statistically non-significant and imprecise, with wide confidence intervals reflecting the limited number of events within these genetic subgroups.

### Predictive Performance and Calibration

We assessed the incremental value of adding sarcopenia to the baseline FRAX model (**Table 4**). The 10-year time-dependent AUC for FRAX Only was 0.71 (95% CI: 0.66–0.77) for MOF and 0.69 (95% CI: 0.60–0.77) for hip fracture. The inclusion of the probable or possible sarcopenia did not increase discriminative power, with AUC values remaining nearly identical across both diagnostic frameworks.

**Table 4.**
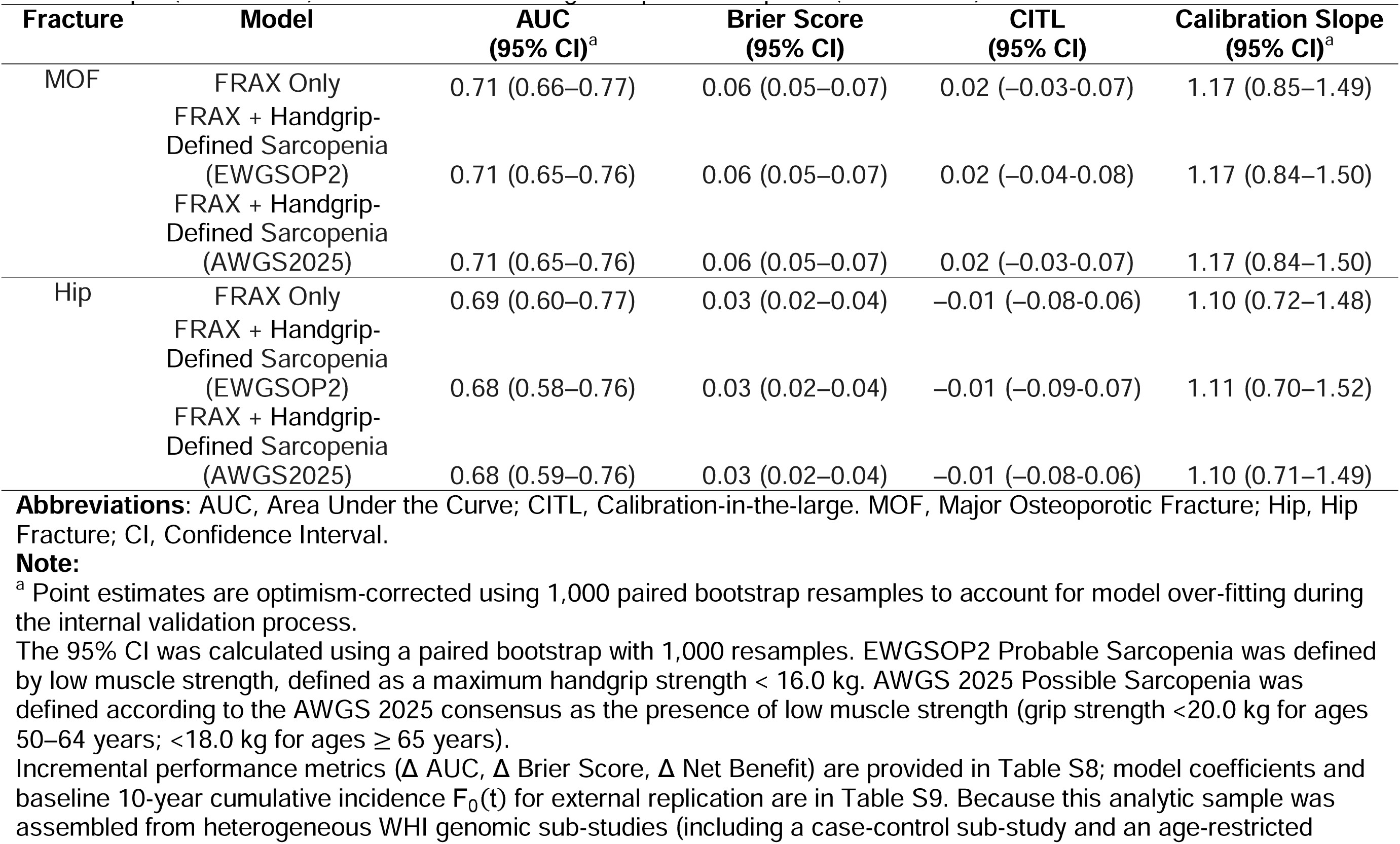
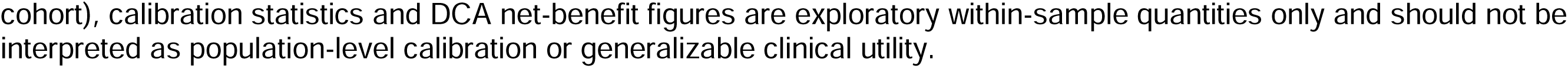
Optimism-Corrected Performance Metrics (Time-Dependent AUC and Calibration) for Major Osteoporotic and Hip Fractures When the Probable or Possible Sarcopenia is Added to the FRAX Model (Without BMD) (n=2,051). Probable/Possible Sarcopenia was diagnosed according to the European Working Group on Sarcopenia in Older People (EWGSOP2) and the Asian Working Group on Sarcopenia (AWGS2025) Criteria.

Calibration metrics indicated adequate internal consistency; Brier scores were consistently low (0.06 for MOF; 0.03 for hip fractures). Calibration-in-the-large (CITL) and calibration slopes (1.17 for MOF; 1.10–1.11 for hip) suggested strong agreement between predicted and observed incidence for the MOF (**Figure 2A**) and hip fracture (**Figure S1A**). Decision curve analysis (**Figure 2B** and **Figure S1B**) corroborated these findings, as the net benefit curves for the sarcopenia-augmented models were indistinguishable from the FRAX-only model across clinically relevant threshold probabilities.

**Figure 2.**
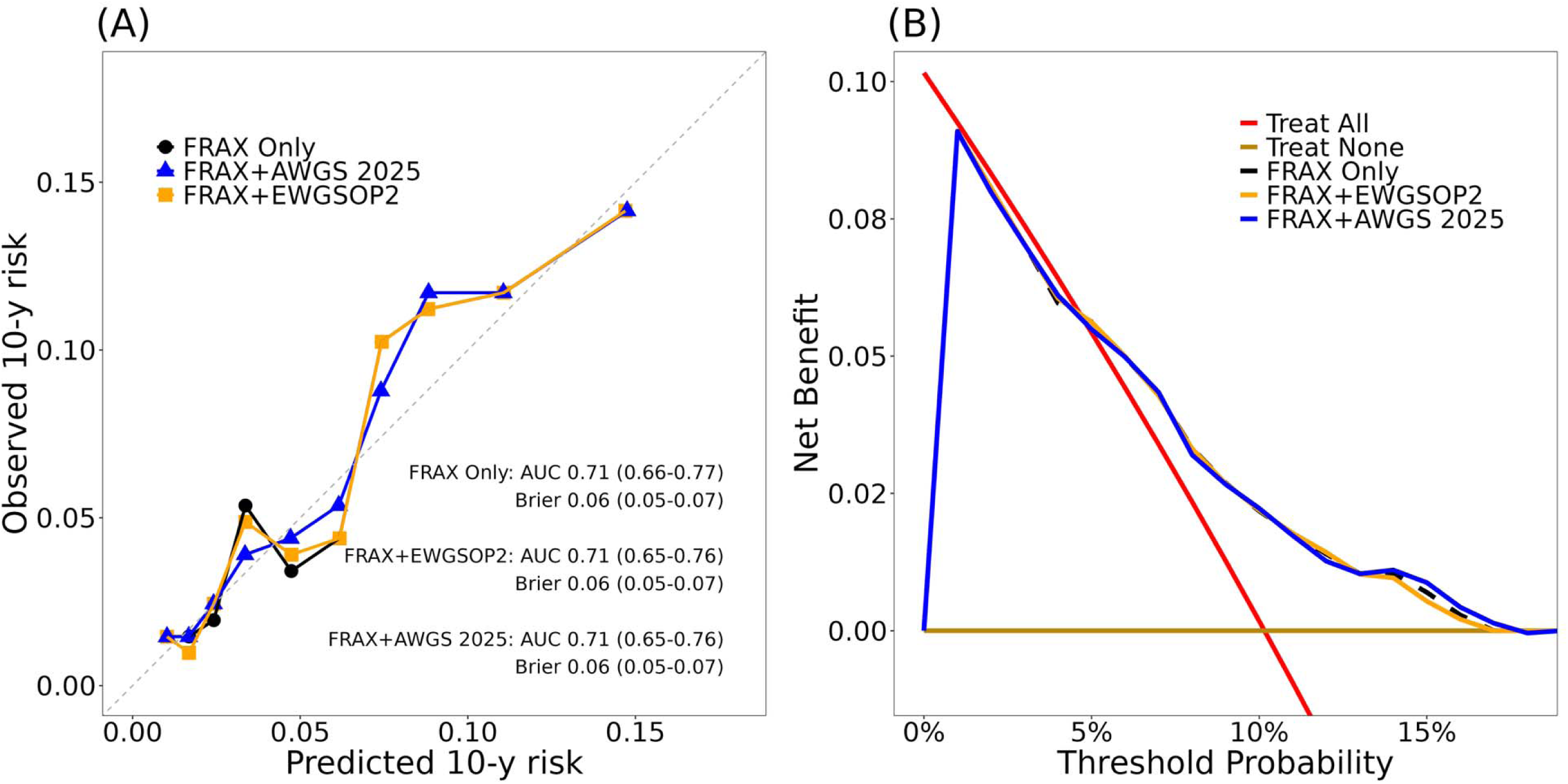
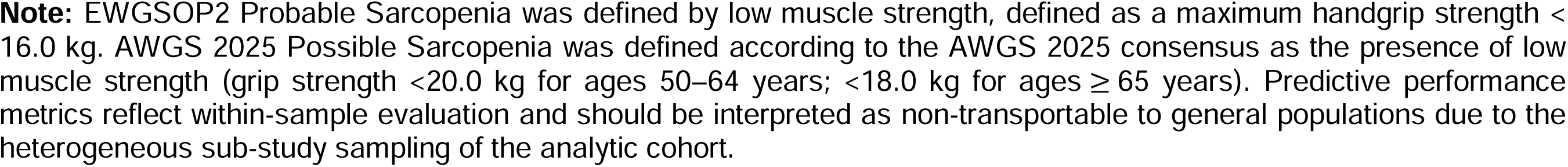
Calibration and decision curve analysis for 10-year major osteoporotic fracture (MOF) risk (n=2,051). **(A)** Calibration plot comparing observed vs. predicted 10-year MOF risk for the FRAX-only model (black), FRAX + EWGSOP2 probable sarcopenia (orange), and FRAX + AWGS 2025 possible sarcopenia (blue). Points represent deciles of predicted risk; the dashed line represents perfect calibration. Text insets display optimism-corrected the time-dependent AUC and Brier score with 95% confidence intervals (CI) calculated via 1,000 bootstrap resamples. **(B)** Decision curve analysis evaluating the net clinical benefit of the models. The x-axis represents the threshold probability for initiating pharmacological intervention, with the clinically relevant range defined as 5% to 20%. The overlapping curves demonstrate a zero change in net benefit (Δ Net Benefit = 0) following the addition of sarcopenia indicators to the baseline FRAX tool.

The performance of the sarcopenia-augmented models was comparable to the FRAX-only model. Formal comparison of the models (**Table S8**) revealed no significant incremental improvement in discrimination (Δ AUC: -0.01 to 0.00) or accuracy (Δ Brier score: -0.001 to 0.000). Furthermore, there was no measurable incremental net clinical benefit within this sample at the designated treatment thresholds (20% for MOF; 3% for Hip), indicating that, within this selected WHI genomic sub-sample, muscle status did not reclassify a clinically meaningful proportion of participants beyond the existing clinical FRAX tool.

### Sensitivity Analysis

To evaluate the robustness of fracture associations outside of the FRAX framework, we performed sensitivity analyses using multivariable models adjusted for age, height, and weight (**Tables 3** and **S6-S7**). These analyses yielded hazard ratios for race/ethnicity and GPS that were consistent in both magnitude and direction with the primary FRAX-adjusted models. To maintain analytic power in this cohort (n=2,051), multiple imputation was employed to address negligible missingness in age (n=2), height (n=3), and weight (n=5); these specifically imputed variables were utilized exclusively within these sensitivity models to ensure the findings remained robust across different adjustment strategies.

### Model Parameters and Internal Validation

This study serves as a model-update and internal validation analysis. To ensure the reproducibility of our sarcopenia-augmented frameworks, we have provided the mathematical parameters, including model coefficients (log-subdistribution hazards) (**Table S9**). As expected in internal validation, these optimism-corrected sHRs show slight shrinkage toward the null compared to the apparent estimates in **Table 3**, and confidence intervals were derived from the same 1,000 bootstrap resamples.

To account for potential over-fitting inherent in model updates, all performance metrics in **Table 4** were adjusted using a bootstrap optimism-correction procedure (1,000 resamples). The optimism-corrected AUC for the MOF outcome remained stable at 0.71, while the corrected calibration slopes (range: 1.10–1.17) indicated minimal over-fitting. These findings confirm that while the addition of sarcopenia did not shift the FRAX discriminative threshold, the internally validated models remain statistically robust and well-calibrated for the 10-year prediction horizon.

For the MOF outcome, the 10-year baseline risk (F_0_(t)) was 0.003, with a sarcopenia coefficient (β) of -0.17 (sHR 0.84) for EWGSOP2 and 0.02 (sHR 1.02) for AWGS 2025. For hip fractures, the baseline risk was approximately 0.008, with a sarcopenia coefficient (β) of 0.06 (sHR 1.06) for the AWGS 2025 augmented model.

While **Table 3** presents the apparent associations within the analytic cohort, the parameters in **Table S9** reflect the optimism-corrected coefficients. As expected in internal validation, the corrected sHRs (e.g., 1.02 for AWGS 2025 MOF) show slight shrinkage toward the null compared to the apparent estimates (e.g., 1.22), ensuring that the updated model is not over-calibrated to the source data.

## Discussion

In this cohort of Black, Hispanic, and White postmenopausal women, our study demonstrates that while handgrip-defined probable or possible sarcopenia and GPS were directionally associated with fracture incidence, neither offered incremental predictive value beyond traditional risk factors already captured by the FRAX tool. Notably, the association between probable or possible sarcopenia status and both MOF and hip fracture lost statistical significance after adjustment for FRAX components. This lack of incremental prognostic value was further supported by time-dependent predictive performance metrics, which revealed no significant improvement in model discrimination (AUC), overall error (Brier score), or within-sample prediction performance (DCA) when handgrip-defined probable or possible sarcopenia was added to the clinical FRAX algorithm. These findings remained robust regardless of the clinical consensus definition employed, including both the EWGSOP2 and AWGS 2025 criteria.

Our results are consistent with a growing body of evidence indicating that the inclusion of consensus-defined sarcopenia does not provide a significant independent contribution to fracture risk prediction in postmenopausal women [31,32]. Our study extends these observations to a broader, Black, Hispanic, and White postmenopausal women US population, suggesting that age-related muscle decline may not contribute substantial unique information to fracture prediction beyond what is already captured by the clinical risk factors within the FRAX tool. This lack of prognostic association in older postmenopausal women (≥65 years) may be explained by the plateauing of muscle health decline relative to other dominant risk factors at this life stage [33].

The lack of a prognostic association between probable or possible sarcopenia and fracture in our adjusted models align with several landmark studies. Schaap et al. [34] found that while individual components of sarcopenia (such as low grip strength) were associated with falls, the consolidated EWGSOP2 definition did not independently predict fractures after adjusting for age and other covariates. Similarly, Cawthon et al. [35] reported that consensus definitions were not superior to simple measurements of walking speed in predicting clinical outcomes.

This lack of incremental prognostic value may be explained by the biological and statistical overlap between sarcopenia and frailty. In our cohort, women with probable or possible sarcopenia were older and had lower body weight and height, factors already heavily weighted in the FRAX algorithm. Consequently, probable or possible sarcopenia may act as a surrogate marker for advanced aging and physical frailty rather than a distinct pathological driver of fracture risk that necessitates separate inclusion in clinical tools.

Our identified racial disparities corroborate literature showing differential calibration of fracture risk tools across ethnic groups. Black women exhibited lower fracture risks than White women, remaining robust after accounting for muscle status. Traditional risk factors and muscle strength may operate differently across racial backgrounds. The lack of incremental predictive value suggests baseline FRAX components, like age and body weight, already capture the effects of musculoskeletal frailty. While the high GPS group (top 5%) showed a higher point estimate for MOF hazard than the low GPS group, differences were non-significant. Adding sarcopenia status did not alter genetic hazard ratios, suggesting genetic risk and muscle strength represent distinct pathways. Within this sample, neither metric improved prediction beyond FRAX; however, small stratum-specific event counts preclude reliable heterogeneity detection, leaving these subgroup results hypothesis-generating.

### Clinical and Public Health Implications

Our results do not support the mandatory inclusion of a sarcopenia diagnosis in standard fracture risk prediction tools. While muscle health is a critical indicator of general health and falls risk, clinicians may find that traditional clinical risk factors provide sufficient discrimination for fracture prediction. Within this exploratory within-sample analysis, DCA showed no measurable incremental net clinical benefit from adding handgrip-defined sarcopenia to the FRAX algorithm across the clinically relevant threshold range. External validation in a prospective cohort is required before drawing clinical utility conclusions.

Our study has several limitations. First, an extremely small sample size and near-absence of fracture events in “Confirmed” or “Severe” sarcopenia subgroups precluded stable statistical analyses; findings are thus limited to strength-based probable or possible stages. Second, baseline data came from an older WHI subset with a higher fracture burden, potentially limiting generalizability. Third, small sample sizes in certain minority groups resulted in wide confidence intervals and reduced statistical power for interaction tests. Fourth, the GPS was derived primarily from European-ancestry populations, which may attenuate precision in Black and Hispanic subgroups. Finally, heterogeneous sampling compromises the transportability of our predictive findings. Including a case-control sub-study (GARNET) and an age-restricted cohort (WHIMS) influenced absolute-risk, calibration, and net-benefit estimates. While maximizing sample size for prognostic evaluation, these outputs remain exploratory within-sample quantities rather than population-level clinical utility. External validation in prospective, population-based cohorts is required.

In a cohort of Black, Hispanic, and White postmenopausal women, a diagnosis of handgrip-defined probable or possible sarcopenia did not provide independent predictive value for 10-year fracture risk beyond established FRAX clinical risk factors within this selected WHI genomic sub-sample. While muscle strength remains vital for assessing physical function, its integration into fracture-specific algorithms to FRAX may not be necessary for improving risk stratification in this population.

## Data Sharing Statement

The data used in the current study is available through controlled access of the database of Genotypes and Phenotypes (dbGap) (https://www.ncbi.nlm.nih.gov/projects/gap/cgi-bin/study.cgi?study_id=phs000200.v12.p3). The summary statistics of the Genome-Wide Association Study used in the current study are available from the Genetic Factors for Osteoporosis Consortium at http://www.gefos.org/?q=content/data-release-2018

## Author contributions

Conceptualization: Jongyun Jung.

Data curation: Jongyun Jung.

Formal analysis: Jongyun Jung.

Methodology: Jongyun Jung.

Software: Jongyun Jung.

Visualization: Jongyun Jung.

Validation: Jongyun Jung.

Writing–original draft: Jongyun Jung.

Writing–review & editing: Qing Wu, Jongyun Jung.

Funding acquisition: Qing Wu.

Project administration: Qing Wu.

Supervision: Qing Wu.

All authors have read and agreed to the published version of the manuscript.

