## Supplemental File for "Prognostic Association of Handgrip-Defined Probable or Possible Sarcopenia Status and Polygenic Risk with 10-Year Fracture Incidence among Black, Hispanic, and White Women: A Women’s Health Initiative Study"

### Supplementary Material

**Table S1. Overview of Women’s Health Initiative Sub-Studies Included in This Analysis.**

| **Study Name** | **dbGaP* Study Accession Number** | **Characteristic of Study** | **Original Sample Size** | **Analytic Sample (n=2,051)** |
| --- | --- | --- | --- | --- |
| Genomics and Randomized Trials Network | phs000315 | - Case-control study within the Genomics and Randomized Trials Network funded by NHGRI - Participants were selected as a nested case-control sample of coronary heart disease, stroke, venous thrombosis, and incident diabetes events. | 4,894 | 501 |
| SNP Health Association Resource | phs000386 | - Prospective cohort study aimed at discovering or replicating genes associated with quantitative traits (e.g., blood pressure, lipids) among African American and Hispanic women. - Conducted with WHI participants consenting to Supplemental Consent for broad data sharing. | 12,007 | 177 |
| Population Architecture using Genomics and Epidemiology | phs000227 | - Prospective cohort study of genotypes generated via BeadXpress and Metabohip, as part of NHGRI's PAGE project. - Eligibility included postmenopausal women aged 50-79, residing in study areas for at least 3 years post-enrollment. | 12,646 | 345 |
| Women’s Health Initiative Memory Study | phs000675 | - Prospective cohort of women aged 65 and older, recruited from the WHI Hormone Trial. - Participants selected for this sub-study include the following WHI Hormone Trial European American women with the appropriate consent for data to be shared on dbGaP. | 5,740 | 733 |
| WHI Sequencing Project | phs000281 | - Prospective cohort using next-generation sequencing to identify genes and mechanisms in heart, lung, and blood disorders. - Eligible participants provided DNA samples and consented to data submission to dbGaP. | 1,904 | 65 |
| WHI Harmonized and Imputed GWAS Data of Hip Fracture | phs000746 | - Prospective study identifying genetic contributors to hip fracture risk through GWAS among postmenopausal WHI participants. | 3,688 | 230 |

*dbGaP: the database of Genotype and Phenotype.

**Table S2**. **Baseline Characteristics of Participants Between the Final Analytic Cohort (n=2,051) and the Excluded (n=25,461) in the Women’s Health Initiative Study (n=27,512).**

| **Variable** | **Final Analytic Cohort**  **(n=2,051)** | **Excluded**  **(n=25,461)** | **SMD** |
| --- | --- | --- | --- |
| Age (years), mean (SD) | 70.1 (3.8) | 63.2 (7.5) | **1.17** |
| Height (cm), mean (SD) | 160.5 (6.0) | 161.1 (6.4) | 0.09 |
| Weight (kg), mean (SD) | 74.6 (15.3) | 76.9 (17.1) | **0.14** |
| FRAX^®^ (without BMD) for MOF (%), mean (SD) | 9.6 (7.8) | 8.2 (7.4) | 0.08 |
| FRAX^®^ (without BMD) for Hip (%), mean (SD) | 2.7 (4.6) | 2.1 (4.1) | 0.05 |
| GPS ^a^, mean (SD) | 4.9 (3.2) | 6.5 (2.8) | **0.48** |
| Self-Reported Race/Ethnicity, $n (\%)$  American Indian or Alaskan Native | 10 (0.5) | 581 (2.3) | **0.72** |
| Asian or Pacific Islander | 32 (1.6) | 467 (1.8) |  |
| Black | 380 (18.5) | 9,675 (38.0) |  |
| Hispanic | 129 (6.3) | 4,505 (17.7) |  |
| White | 1,500 (73.1) | 10,233 (40.2) |  |
| MOF, $n (\%)$ | 134 (6.5) | 1,290 (5.1) | 0.06 |
| Hip Fracture, $n (\%)$ | 73 (3.6) | 845 (3.3) | 0.01 |
| Current Smoking, $n (\%)$ | 132 (6.4) | 2,447 (9.6) | **0.12** |
| Rheumatoid Arthritis, $n (\%)$ | 240 (11.7) | 3,431 (13.5) | 0.05 |
| Previous fragility fracture, $n (\%)$ | 35 (1.7) | 353 (1.4) | 0.03 |
| Parental hip fracture history, $n (\%)$ | 281 (13.7) | 2,503 (9.8) | **0.12** |
| Alcohol intake, $n (\%)$ | 1,419 (69.2) | 16,588 (65.2) | 0.09 |
| ≥ 1 Previous falls in the past 12 months, $n (\%)$ | 682 (33.3) | 8,282 (32.5) | 0.02 |

**Abbreviations**: SD, Standard Deviation; BMD, Bone Mineral Density; GPS, Genome-Wide Polygenic Score; MOF, Major Osteoporotic Fractures, SMD, Standardized Mean Difference.

Boldface denotes meaningful imbalances in SMD (>0.10).

^a^ Calculated using the Genome-Wide Association Study Summary Statistics from the UK Biobank involving 103,155 genetic variants [16].

**Table S3. Baseline Characteristics of Participants in the Women’s Health Initiative Study Stratified by the Asian Working Group for Sarcopenia 2025 Consensus Update (AWGS 2025) Possible Sarcopenia Status (n=2,051).**

| **Variable** | **Normal**  **(n=1,919)** | **Possible Sarcopenia**  **(n=132)** | **P-value*** | **SMD** |
| --- | --- | --- | --- | --- |
| Age (years), mean (SD) | 70.1 (3.8) | 70.5 (3.9) | 0.17 | **0.12** |
| Height (cm), mean (SD) | 160.7 (6.0) | 158.1 (5.9) | **<0.01** | **0.43** |
| Weight (kg), mean (SD) | 74.9 (15.4) | 70.4 (13.7) | **<0.01** | **0.31** |
| FRAX^®^ (without BMD) for MOF (%), mean (SD) | 9.5 (7.8) | 9.8 (8.1) | 0.64 | 0.04 |
| FRAX^®^ (without BMD) for Hip (%), mean (SD) | 2.7 (4.6) | 2.8 (4.4) | 0.79 | 0.03 |
| GPS ^a^, mean (SD) | 5.0 (2.9) | 5.2 (2.9) | 0.49 | 0.06 |
| Self-Reported Race/Ethnicity, $n (\%)$  American Indian or Alaskan Native | 10 (0.5) | 0 (0.0) | 0.15 | **0.25** |
| Asian or Pacific Islander | 30 (1.6) | 2 (1.5) |  |  |
| Black | 364 (19.0) | 16 (12.1) |  |  |
| Hispanic | 116 (6.0) | 13 (9.8) |  |  |
| White | 1,399 (72.9) | 101 (76.5) |  |  |
| MOF, $n (\%)$ | 123 (6.4) | 11 (8.3) | 0.34 | **0.12** |
| Hip Fracture, $n (\%)$ | 68 (3.5) | 5 (3.8) | 0.62 | 0.08 |
| Current Smoking, $n (\%)$ | 123 (6.4) | 9 (6.8) | 0.91 | 0.02 |
| Rheumatoid Arthritis, $n (\%)$ | 217 (11.3) | 23 (17.4) | 0.05 | **0.18** |
| Previous fragility fracture, $n (\%)$ | 28 (1.5) | 7 (5.3) | **<0.01** | **0.21** |
| Parental hip fracture history, $n (\%)$ | 270 (14.1) | 11 (8.3) | 0.09 | **0.18** |
| Alcohol intake, $n (\%)$ | 1,331 (69.4) | 88 (66.7) | 0.58 | 0.06 |
| ≥ 1 Previous falls in the past 12 months, $n (\%)$ | 637 (33.2) | 45 (34.1) | 0.91 | 0.02 |

**Abbreviations**: SD, Standard Deviation; GPS, Genome-Wide Polygenic Score; MOF, Major Osteoporotic Fractures; SMD, Standardized Mean Difference.

**Note:** Possible Sarcopenia was defined according to the AWGS 2025 consensus as the presence of low muscle strength (grip strength <20.0 kg for ages 50–64 years; <18.0 kg for ages $\geq$65 years).

* P-value obtained by t-test for continuous variables and chi-square tests (or Fisher's Exact Test where appropriate) for categorical variables.

Boldface denotes statistically significant p-values (<0.05) and meaningful imbalances in SMD (>0.10).

^a^ Calculated using the Genome-Wide Association Study Summary Statistics from the UK Biobank involving 103,155 genetic variants [16].

**Table S4**. **10-Year Cumulative Incidence of Major Osteoporotic and Hip Fractures Stratified by Self-Reported Race/Ethnicity and Probable or Possible Sarcopenia Status According to the European Working Group on Sarcopenia in Older People (EWGSOP2) and the Asian Working Group for Sarcopenia 2025 Consensus Update (AWGS 2025) Criteria. Analytical sample includes Black, Hispanic, and White participants (n=2,009).**

| **Fracture** | **Race** | **Sarcopenia Status** | **EWGSOP2** | | |  | **AWGS 2025** | | |
| --- | --- | --- | --- | --- | --- | --- | --- | --- | --- |
|  |  |  | **n** | **Events** | **10-Year Cumulative Incidence (95% CI)** |  | **n** | **Events** | **10-Year Cumulative Incidence (95% CI)** |
| MOF | Black (n=380) | Normal | 365 | 4 | 1.6% (0.5% - 3.9%) |  | 364 | 5 | 2.0% (0.8% - 4.4%) |
|  |  | Probable/Possible | 15 | 1 | 10.1% (0.4% - 38.1%) |  | 16 | 0 | 0% (NA) |
|  | Hispanic (n=129) | Normal | 118 | 7 | 9.0% (3.9% - 16.7%) |  | 116 | 6 | 7.9% (3.2% - 15.5%) |
|  |  | Probable/Possible | 11 | 1 | 11.1% (0.5% - 40.6%) |  | 13 | 2 | 19.2% (2.5% - 47.6%) |
|  | White (n=1,500) | Normal | 1,435 | 114 | 10.6% (8.9% - 12.6%) |  | 1,399 | 109 | 10.5% (8.7% - 12.4%) |
|  |  | Probable/Possible | 65 | 4 | 8.1% (2.5% - 17.9%) |  | 101 | 9 | 11.3% (5.5% - 19.5%) |
| Hip | Black (n=380) | Normal | 365 | 1 | 0.4% (0% - 2.2%) |  | 364 | 1 | 0.4% (0% - 2.2%) |
|  |  | Probable/Possible | 15 | 0 | 0% (NA) |  | 16 | 0 | 0% (NA) |
|  | Hispanic (n=129) | Normal | 118 | 1 | 1.5% (0.1% - 7.4%) |  | 116 | 1 | 1.5% (0.1% - 7.4%) |
|  |  | Probable/Possible | 11 | 0 | 0% (NA) |  | 13 | 0 | 0% (NA) |
|  | White (n=1,500) | Normal | 1,435 | 64 | 6.3% (4.9% - 7.9%) |  | 1,399 | 63 | 6.3% (4.9% - 7.9%) |
|  |  | Probable/Possible | 65 | 4 | 8.1% (2.5% - 17.9%) |  | 101 | 5 | 6.8% (2.5% - 14.2%) |

**Abbreviations:** MOF, Major Osteoporotic Fractures; Hip, Hip Fracture.

**Note**: The total cohort size was n=2,051. American Indian or Alaskan Native (n=10) and Asian or Pacific Islander (n=32) individuals were excluded due to the low incidence of fractures during follow-up. The 10-Year Cumulative Incidence values represent the estimated 10-year probability of the fractures. These estimates were calculated using the Aalen-Johansen estimator to account for the competing risk of mortality and right-censoring. This approach ensures that the reported risk is not overestimated by ignoring those who died before experiencing a fracture. EWGSOP2 Probable Sarcopenia was defined by low muscle strength, defined as a maximum handgrip strength < 16.0 kg. AWGS 2025 Possible Sarcopenia was defined according to the AWGS 2025 consensus as the presence of low muscle strength (grip strength <20.0 kg for ages 50–64 years; <18.0 kg for ages $\geq$65 years). Strata with zero events (shown as 0% [NA]) have insufficient data to estimate 10-year cumulative incidence or support interaction tests; estimates from these strata should be interpreted with extreme caution.

**Table S5**. **10-Year Cumulative Incidence of Major Osteoporotic and Hip Fractures Stratified by the Genome-Wide Polygenic Score (GPS) and Probable or Possible Sarcopenia Status According to the European Working Group on Sarcopenia in Older People (EWGSOP2) and the Asian Working Group for Sarcopenia 2025 Consensus Update (AWGS 2025) Criteria (n=2,051).**

| **Outcome** | **GPS** | **Sarcopenia** | **EWGSOP2** | | |  | **AWGS 2025** | | |
| --- | --- | --- | --- | --- | --- | --- | --- | --- | --- |
|  |  |  | **n** | **Events** | **10-Year Cumulative Incidence (95% CI)** |  | **n** | **Events** | **10-Year Cumulative Incidence (95% CI)** |
| MOF | High (n=103) | Normal | 99 | 11 | 13.7% (7.2% - 22.3%) |  | 96 | 11 | 14.1% (7.4% - 22.8%) |
|  |  | Probable/Possible | 4 | 0 | 0% (NA) |  | 7 | 0 | 0% (NA) |
|  | Medium (n=1,845) | Normal | 1,761 | 112 | 8.8% (7.3% - 10.5%) |  | 1,725 | 107 | 8.6% (7.1% - 10.3%) |
|  |  | Probable/Possible | 84 | 6 | 9.7% (3.9% - 18.7%) |  | 120 | 11 | 12.0% (6.3% - 19.7%) |
|  | Low (n=103) | Normal | 100 | 5 | 6.8% (2.5% - 14.1%) |  | 98 | 5 | 6.9% (2.5% - 14.3%) |
|  |  | Probable/Possible | 3 | 0 | 0% (NA) |  | 5 | 0 | 0% (NA) |
| Hip | High (n=103) | Normal | 99 | 4 | 5.6% (1.8% - 12.7%) |  | 96 | 4 | 5.8% (1.8% - 13.1%) |
|  |  | Probable/Possible | 4 | 0 | 0% (NA) |  | 7 | 0 | 0% (NA) |
|  | Medium (n=1,845) | Normal | 1,761 | 62 | 5.1% (3.9% - 6.4%) |  | 1,725 | 61 | 5.1% (3.9% - 6.4%) |
|  |  | Probable/Possible | 84 | 4 | 6.7% (2.1% - 15.1%) |  | 120 | 5 | 6.0% (2.2% - 12.6%) |
|  | Low (n=103) | Normal | 100 | 3 | 4.2% (1.1% - 10.6%) |  | 98 | 3 | 4.2% (1.1% - 10.8%) |
|  |  | Probable/Possible | 3 | 0 | 0% (NA) |  | 5 | 0 | 0% (NA) |

**Abbreviations:** MOF, Major Osteoporotic Fractures; Hip, Hip Fracture.

**Note**: GPS categories were defined as follows: low (bottom 5%), medium (middle 90%), and high (top 5%). The 10-Year Cumulative Incidence values represent the estimated 10-year probability of the fractures. These estimates were calculated using the Aalen-Johansen estimator to account for the competing risk of mortality and right-censoring. This approach ensures that the reported risk is not overestimated by ignoring those who died before experiencing a fracture. EWGSOP2 Probable Sarcopenia was defined by low muscle strength, defined as a maximum handgrip strength < 16.0 kg. AWGS 2025 Possible Sarcopenia was defined according to the AWGS 2025 consensus as the presence of low muscle strength (grip strength <20.0 kg for ages 50–64 years; <18.0 kg for ages $\geq$65 years). Strata with zero events (shown as 0% [NA]) have insufficient data to estimate 10-year cumulative incidence or support interaction tests; estimates from these strata should be interpreted with extreme caution.

**Table S6**. **Self-Reported Race/Ethnicity Differences in Risk of Major Osteoporotic Fracture (MOF) and Hip Fracture in the Women’s Health Initiative Study**. Handgrip-Defined Probable or Possible Sarcopenia was ascertained according to the European Working Group on Sarcopenia in Older People (EWGSOP2) and the Asian Working Group for Sarcopenia 2025 Consensus Update (AWGS 2025) Criteria. Analytical sample includes Black, Hispanic, and White participants (n=2,009).

| **Fracture** | **Model** | **Self-Reported Race/Ethnicity** | **EWGSOP2 sHR (95% CI)** | **AWGS 2025 sHR (95% CI)** |
| --- | --- | --- | --- | --- |
| MOF | FRAX Only | White (n=1,500) | 1.00 (Reference) | 1.00 (Reference) |
|  |  | Black (n=380) | **0.22 (0.09–0.53)** | **0.22 (0.09–0.53)** |
|  |  | Hispanic (n=129) | 1.06 (0.52–2.17)* | 1.06 (0.52–2.17)* |
|  | FRAX + Handgrip-Defined Probable/Possible Sarcopenia | White (n=1,500) | 1.00 (Reference) | 1.00 (Reference) |
|  |  | Black (n=380) | **0.22 (0.09–0.53)** | **0.22 (0.09–0.53)** |
|  |  | Hispanic (n=129) | 1.06 (0.52–2.17)* | 1.05 (0.51–2.15)* |
|  | Adjusted** + Handgrip-Defined Probable/Possible Sarcopenia | White (n=1,500) | 1.00 (Reference) | 1.00 (Reference) |
|  |  | Black (n=380) | **0.19 (0.08–0.48)** | **0.19 (0.08–0.48)** |
|  |  | Hispanic (n=129) | 1.00 (0.47–2.14)* | 1.00 (0.48–2.08)* |
| Hip | FRAX Only | White (n=1,500) | 1.00 (Reference) | 1.00 (Reference) |
|  |  | Black (n=380) | **0.07 (0.01–0.48)** | **0.07 (0.01–0.48)** |
|  |  | Hispanic (n=129) | 0.21 (0.03–1.51)* | 0.21 (0.03–1.51)* |
|  | FRAX + Handgrip-Defined Probable/Possible Sarcopenia | White (n=1,500) | 1.00 (Reference) | 1.00 (Reference) |
|  |  | Black (n=380) | **0.07 (0.01–0.48)** | **0.07 (0.01–0.48)** |
|  |  | Hispanic (n=129) | 0.21 (0.03–1.50)* | 0.21 (0.03–1.50)* |
|  | Adjusted** + Handgrip-Defined Probable/Possible Sarcopenia | White (n=1,500) | 1.00 (Reference) | 1.00 (Reference) |
|  |  | Black (n=380) | **0.07 (0.01–0.52)** | **0.07 (0.01–0.52)** |
|  |  | Hispanic (n=129) | 0.31 (0.04–2.19)* | 0.31 (0.04–2.19)* |

**Abbreviations:** sHR, Subdistribution Hazard Ratio comparing indicated group to the "White" reference; CI, Confidence Interval. MOF, Major Osteoporotic Fractures; Hip, Hip Fracture.

**Note**: The total cohort size was n=2,051. American Indian or Alaskan Native (n=10) and Asian or Pacific Islander (n=32) individuals were excluded due to the low incidence of fractures during follow-up. Significant results are in boldface. FRAX Score without BMD was used in all analysis. FRAX hip fracture risk score was used in the hip fracture models. EWGSOP2 Probable Sarcopenia was defined by low muscle strength, defined as a maximum handgrip strength < 16.0 kg. AWGS 2025 Possible Sarcopenia was defined according to the AWGS 2025 consensus as the presence of low muscle strength (grip strength <20.0 kg for ages 50–64 years; <18.0 kg for ages $\geq$65 years).

* These hazard ratios and confidence intervals are likely influenced by small subgroup sizes or low event counts within the specific strata, resulting in high standard errors and wide confidence intervals.

** Covariates included: age, height, and weight.

Because this sample was drawn from heterogeneous WHI genomic sub-studies, stratum-specific sHRs are exploratory within-sample estimates and should not be interpreted as transportable population-level associations.

**Table S7**. **Genome-Wide Polygenic Score (GPS) Group Differences in Risk of Major Osteoporotic Fracture (MOF) and Hip Fracture in the Women’s Health Initiative Study (n=2,051).** Handgrip-Defined Probable or Possible Sarcopenia was ascertained according to the European Working Group on Sarcopenia in Older People (EWGSOP2) and the Asian Working Group for Sarcopenia 2025 Consensus Update (AWGS 2025) Criteria.

| **Fracture** | **Model** | **GPS** | **EWGSOP2 sHR (95% CI)** | **AWGS 2025 sHR (95% CI)** |
| --- | --- | --- | --- | --- |
| MOF | FRAX Only | Low (n=103) | 1.00 (Reference) | 1.00 (Reference) |
|  |  | Medium (n=1,845) | 1.30 (0.53–3.19) | 1.30 (0.53–3.19) |
|  |  | High (n=103) | 1.75 (0.61–5.02)* | 1.75 (0.61–5.02)* |
|  | FRAX + Handgrip-Defined Probable/Possible Sarcopenia | Low (n=103) | 1.00 (Reference) | 1.00 (Reference) |
|  |  | Medium (n=1,845) | 1.30 (0.53–3.19) | 1.30 (0.53–3.19) |
|  |  | High (n=103) | 1.75 (0.61–5.02)* | 1.75 (0.61–5.02)* |
|  | Adjusted** + Handgrip-Defined Probable/Possible Sarcopenia | Low (n=103) | 1.00 (Reference) | 1.00 (Reference) |
|  |  | Medium (n=1,845) | 1.49 (0.60–3.69) | 1.49 (0.60–3.69) |
|  |  | High (n=103) | 2.34 (0.81–6.81)* | 2.34 (0.81–6.81)* |
| Hip | FRAX Only | Low (n=103) | 1.00 (Reference) | 1.00 (Reference) |
|  |  | Medium (n=1,845) | 1.26 (0.39–4.03) | 1.26 (0.39–4.03) |
|  |  | High (n=103) | 1.19 (0.27–5.32)* | 1.19 (0.27–5.32)* |
|  | FRAX + Handgrip-Defined Probable/Possible Sarcopenia | Low (n=103) | 1.00 (Reference) | 1.00 (Reference) |
|  |  | Medium (n=1,845) | 1.26 (0.39–4.02) | 1.26 (0.39–4.02) |
|  |  | High (n=103) | 1.19 (0.27–5.31)* | 1.19 (0.27–5.31)* |
|  | Adjusted** + Handgrip-Defined Probable/Possible Sarcopenia | Low (n=103) | 1.00 (Reference) | 1.00 (Reference) |
|  |  | Medium (n=1,845) | 1.38 (0.43–4.49) | 1.38 (0.43–4.49) |
|  |  | High (n=103) | 1.47 (0.32–6.69)* | 1.47 (0.32–6.69)* |

**Abbreviations:** sHR, Subdistribution Hazard Ratio comparing indicated group to the "Low" reference; CI, Confidence Interval. MOF, Major Osteoporotic Fractures; Hip, Hip Fracture.

**Note**: GPS categories were defined as follows: Low (bottom 5%), Medium (middle 90%), and High (top 5%). All models in this table include adjustment for genotyping array, sub-study indicators, and the first 10 genetic principal components. ‘FRAX Only’ in this table therefore denotes the FRAX score plus this genetic adjustment set; ‘FRAX + Handgrip-Defined Sarcopenia’ additionally includes binary sarcopenia status. FRAX Score without BMD was used in all analysis. FRAX hip fracture risk score was used in the hip fracture models. EWGSOP2 Probable Sarcopenia was defined by low muscle strength, defined as a maximum handgrip strength < 16.0 kg. AWGS 2025 Possible Sarcopenia was defined according to the AWGS 2025 consensus as the presence of low muscle strength (grip strength <20.0 kg for ages 50–64 years; <18.0 kg for ages $\geq$65 years).

* These hazard ratios and confidence intervals are likely influenced by small subgroup sizes or low event counts within the specific strata, resulting in high standard errors and wide confidence intervals.

** **Adjusted + Probable or Possible Sarcopenia:** These models include the baseline genetic adjustment set (array, sub-study indicators, and 10 PCs) plus age, height, and weight.

Because this sample was drawn from heterogeneous WHI genomic sub-studies, stratum-specific sHRs are exploratory within-sample estimates and should not be interpreted as transportable population-level associations.

**Table S8.** **Incremental Change in Predictive Performance Metrics Following the Addition of Sarcopenia to the FRAX Framework.**

| **Fracture** | **Model Comparison** | $\text{Δ}$ **AUC (95% CI)^a^** | $\text{Δ}$ **Brier Score (95% CI)** | $\text{Δ}$ **Net Benefit^b^** | **P-value^c^** |
| --- | --- | --- | --- | --- | --- |
| MOF | FRAX + EWGSOP2 vs. FRAX Only | 0.00 (-0.01 to 0.01) | -0.001 (-0.002 to 0.001) | 0 | 0.89 |
|  | FRAX + AWGS 2025 vs. FRAX Only | 0.00 (-0.01 to 0.01) | -0.001 (-0.003 to 0.002) | 0 | 0.92 |
| Hip | FRAX + EWGSOP2 vs. FRAX Only | -0.01 (-0.02 to 0.01) | 0.000 (-0.001 to 0.001) | 0 | 0.45 |
|  | FRAX + AWGS 2025 vs. FRAX Only | -0.01 (-0.02 to 0.01) | 0.000 (-0.001 to 0.001) | 0 | 0.51 |

**Abbreviations**: AUC, Area Under the Curve; MOF, Major Osteoporotic Fracture; Hip, Hip Fracture; CI, Confidence Interval.

**Note:**

^a^ $\text{Δ}$ **Metrics**: Calculated as the metric for the Augmented Model minus the metric for the FRAX Only model. Negative

$\text{Δ}$ Brier scores indicate improved accuracy.

^b^ $\text{Δ}$ **Net Benefit**: Calculated at the clinical treatment threshold of 20% for MOF and 3% for Hip Fracture.

^c^ **P-value**: Derived from 1,000 paired-bootstrap resamples comparing the models.

**Table S9. Model Parameters for Handgrip-Defined Probable or Possible** **Sarcopenia-Augmented FRAX Frameworks (n=2,051).**

| **Outcome** | **Model Framework** | **Baseline 10-y Risk** $\left( \text{F}_{\text{0}}\left( \text{t} \right) \right)$**^a^** | **Sarcopenia sHR (95% CI)^b^** | **Coefficient (**$\text{β)}$^c^ |
| --- | --- | --- | --- | --- |
| MOF | FRAX + EWGSOP2 | 0.003 | 0.84 (0.62–1.13) | -0.176 |
|  | FRAX + AWGS 2025 | 0.003 | 1.02 (0.79–1.30) | 0.015 |
| Hip Fracture | FRAX + EWGSOP2 | 0.008 | 0.85 (0.64–1.14) | -0.158 |
|  | FRAX + AWGS 2025 | 0.008 | 1.06 (0.83–1.34) | 0.055 |

**Abbreviations:** MOF, Major Osteoporotic Fracture; sHR, Subdistribution Hazard Ratio; CI, Confidence Interval.

**Notes:**

^a^ **Baseline 10-year Risk** $\left( \text{F}_{\text{0}}\left( \text{t} \right) \right)$**:** Represents the cumulative incidence of the fracture event at 10 years for a participant with a FRAX score of 0 and a "Normal" muscle status.

^b^ **Subdistribution Hazard Ratio (sHR):** Derived from the Fine-Gray subdistribution models to account for the competing risk of mortality. Point estimates are optimism-corrected via a 1,000-resample bootstrap procedure: for each resample, a model was fit on the bootstrap sample and evaluated on both the bootstrap sample and the original dataset; optimism was computed as the average performance difference. Corrected values differ from apparent sHRs in Table 3 and are intended for external replication, not hypothesis testing.

^c^ **Coefficient (**$\text{β)}$**:** The log-subdistribution hazard ratio (ln(sHR)). This is the actual "mathematical parameter" used in the model equation.

The sHRs and coefficients reported in Table S9 are optimism-corrected via 1,000 bootstrap resamples. These values differ from the apparent sHRs reported in Table 3, as they have been adjusted for potential model over-fitting to provide more robust parameters for model replication and future validation.

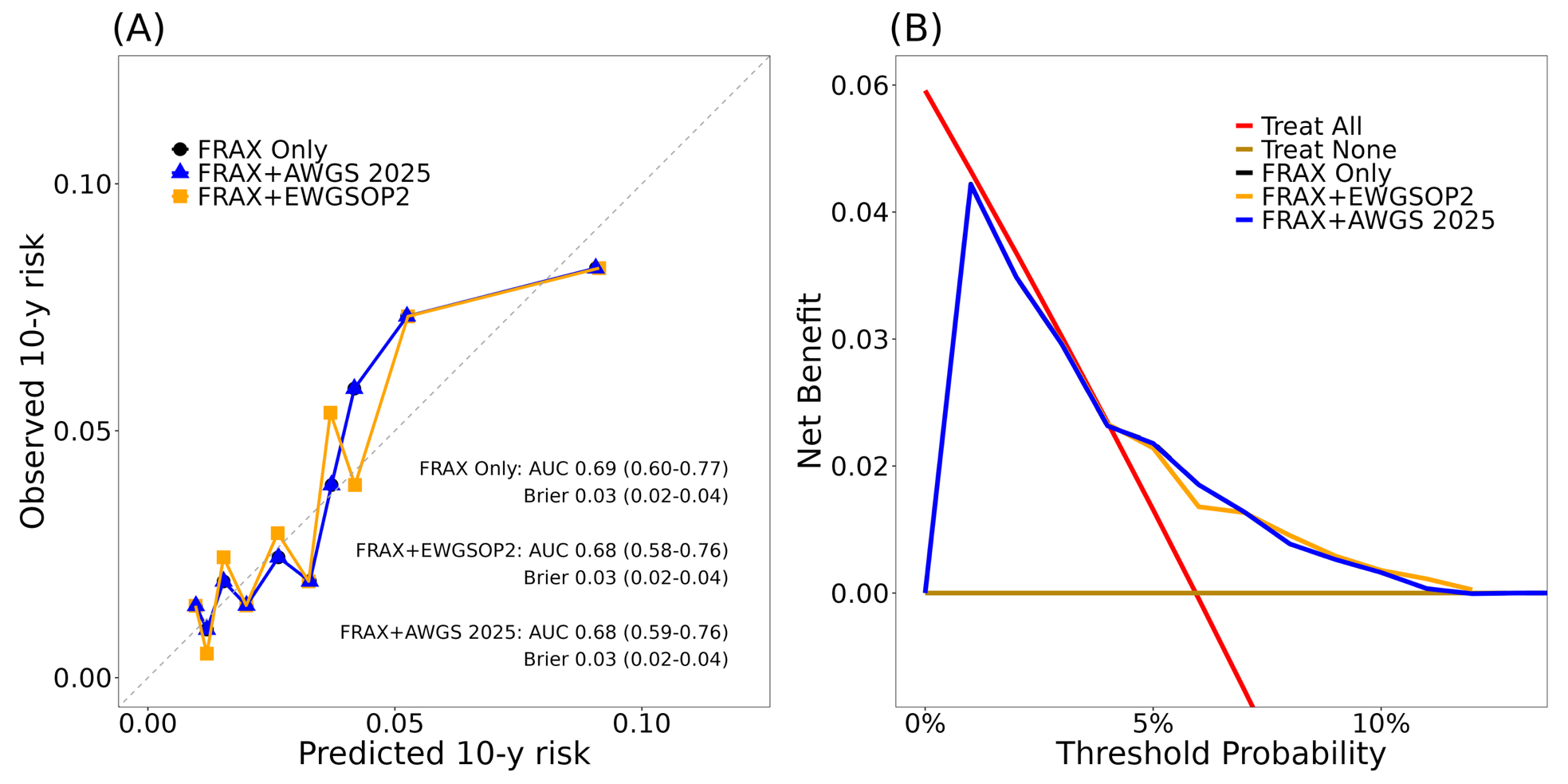
**Figure S1**. **Calibration and decision curve analysis for 10-year hip fracture risk (n=2,051).** **(A)** Calibration plot comparing observed vs. predicted 10-year hip fracture risk for the FRAX-only model (black), FRAX + EWGSOP2 probable sarcopenia (orange), and FRAX + AWGS 2025 possible sarcopenia (blue). Points represent deciles of predicted risk; the dashed line represents perfect calibration. Text insets show optimism-corrected the time-dependent AUC and Brier score with 95% confidence intervals (CI) calculated via 1,000 bootstrap resamples. **(B)** Decision curve analysis illustrating the net clinical benefit. The clinically relevant threshold for hip fracture intervention is highlighted at 3%. As indicated by the absence of curve separation, the augmented models provided no measurable incremental within-sample net benefit ($\text{Δ}$ Net Benefit = 0) over the FRAX-only model at this threshold.

**Note:** EWGSOP2 Probable Sarcopenia was defined by low muscle strength, defined as a maximum handgrip strength < 16.0 kg. AWGS 2025 Possible Sarcopenia was defined according to the AWGS 2025 consensus as the presence of low muscle strength (grip strength <20.0 kg for ages 50–64 years; <18.0 kg for ages $\geq$65 years). Predictive performance metrics reflect within-sample evaluation and should be interpreted as non-transportable to general populations due to the heterogeneous sub-study sampling of the analytic cohort.
